# Influence of parenting styles on depression, anxiety, stress and self-esteem of adolescents

**DOI:** 10.1101/2024.08.16.24312121

**Authors:** Rabina Khadka, Anjali Bhatt, Milan Thapa, Anusha Sharma, Manoj Joshi, Durga Khadka Mishra

## Abstract

Globally, fourteen percent adolescents suffer from mental health issues. Among the many contributing factors to mental health problems, parenting is considered to have a significant effect on both the physical and psychological health of adolescents. This study aimed to assess depression, anxiety, stress, and self-esteem among adolescents and their relationship with parenting style. A community-based cross-sectional study with multistage proportionate sampling was conducted among 583 school-going adolescents from September 2022 to March 2023 in Bheemdatt Municipality. This study used the previously validated Depression Anxiety and Stress Scale-21 (DASS-21), Rosenberg Self-Esteem Scale, and Parenting Style and Dimension Questionnaire to assess the status and relationships. Variables were analyzed using a multivariate logistic regression model, and a p-value of <0.05 was considered statistically significant. The prevalence of depression, anxiety, and stress among adolescents was found to be 37.39% (95% CI: 33.46 to 41.32), 42.19% (95% CI: 38.18 to 46.19), and 24.69% (95% CI: 21.18 to 28.19), respectively. Likewise, the mean self-esteem score among adolescents was found to be 22.26 ± 3.56. The authoritative parenting style was positively associated with DAS, while the authoritarian and permissive parenting styles were negatively associated with DAS and positively associated with self-esteem. Parent’s age, parent’s education, social support, and bullying, showed strong effects on the research outcomes. These findings highlight the crucial role of parental involvement and support in shaping adolescent mental health, underscoring the need for interventions that encourage positive parenting practices to improve mental health outcomes for adolescents. It is highly recommended that school-based programs focus on raising awareness of mental health issues and offer interventions such as stress management and student counseling. There is a pressing need to effectively monitor and enforce anti-bullying policies within educational settings.

## Introduction

“Adolescence is the phase of life between childhood and adulthood, from ages 10 to 19. It is a unique stage of human development and an important time for laying the foundations of good health.”[1] Adolescents experience rapid physical, cognitive, psychological, and social changes, including exposure to poverty, abuse, or violence, which can make adolescents vulnerable to mental health problems. Most of these conditions remain largely unrecognized and untreated.[2] Globally, 1 in 7 (14%) of 10 to 19-year-olds worldwide suffer from mental health issues.[3] As per the National Mental Health Survey (NMHS), Nepal 2020, the prevalence of mental disorders among adolescents was recorded at 5.2%.[4] Moreover, within Sudurpashim province, the prevalence of mental disorders was found to be 3.9%.[4] The issue of mental health is getting worse in Nepal, and adolescents are disproportionately affected.[4–6] Adolescents exposure to risk factors like living conditions, stigma, discrimination, family situation, or lack of access to quality health services, make them vulnerable to mental health conditions.[2] Indeed, parenting style has been found to have a long-lasting effect on the development of teenagers’ personalities and other psychological traits in addition to having a direct impact on their mental health.[7–9]

A survey by WHO in 2017 reported that among the countries of South East Asia, Nepal had the highest rates of suicidal ideation (14%) and a significant number of suicide attempts (10%) among adolescents. Additionally, 5% and 7% of adolescents, showed signs of anxiety and loneliness respectively.[6] Further survey revealed that perceived parental engagement was 54% for “parents understood their problems and worries”, 51% for “parents really knew what they were doing with their free time” and 50% for “parents checked if homework was done”.[6] Adolescents whose parents who were less engaged in their lives had attempted suicide at a rate of 15% and 6% faced anxiety and loneliness respectively.[6] As a result, it is crucial to investigate how parenting style affects adolescents’ mental health.

According to Baumrind 1971, there are three types of parenting styles: authoritative, permissive, and authoritarian.[10] Previous studies have shown that these parenting styles yield varying results on the mental health outcome of a child.[11–14] Parents play a major role in the physical and emotional development of their child. Various research has shown the influence of parenting style on the emotional development of adolescents.[11–13,15,16] Recent studies conducted in Nepal has examined how parents can enhance their adolescent’s self-esteem and prevent suicidal behavior, revealing an association between parental authoritativeness and the self-esteem of their adolescents.[12] Prevalence of suicidal risk behavior among adolescents (11.3%) strongly correlates with authoritarian parents.[12]

Taking into account factors affecting mental health, there is limited research highlighting the impact of parenting style on adolescents’ mental health, more particularly concerning depression, anxiety, stress, and self-eteem.[12] Therefore, this study aimed to assess depression, anxiety, stress, and self-esteem in adolescents and their relationship with parenting style in Bheemdatt Municipality, Kanchanpur, Nepal.

## Materials and methods

### Study setting, design and population

The cross-sectional study was conducted among school-going adolescents of Bheemdatt Municipality in Kanchanpur district of Sudurpaschim Province, Nepal. The study was conducted between mid-September 2022 to March 2023. The municipality was named Mahendranagar in honor of the late King Mahendra of Nepal.[17] Later, it was changed to Bhimdatta municipality in honor of the revolutionary farmer leader Bhimdatta Panta, following the country’s 2008 republican transition.[17] The municipality has 19 wards with the area of 171.24 km^2^ (66.12 sq mi) and located 700 kilometers (430 miles) west of Kathmandu and 5 kilometers (3.1 mi) east of the Indian border.[17] Data collection was done in the first week of September 2022. There were 124 schools in the municipality with a population of 5648, out of which 10 schools were selected. School-going adolescents were the study population. These school-going adolescents comprised of the adolescents of grade 9 and 10.

### Sample size calculation and sampling technique

A multistage proportionate sampling technique was used to select the schools and participants randomly. The sample size was estimated using the formula for cross-sectional survey 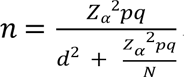using p = 0.57 and q = 0.43 [18] and finite population of 5648 adolescents at confidence interval of 95% and allowable error set at 5%. After the calculation, minimum sample size was 353. After adjusting design effect of 1.5 and adjusting variance from cluster design and assuming nonresponse at 10%, the final sample size was 583. Two-stage cluster sampling was used where two separate lists of public and private schools of Bheemdatt Municipality were prepared and then, ten schools (5 public and 5 private schools) were chosen from the list of schools that fall under the criteria. Schools with less than 50 students were excluded from the study. Likewise, adolescents without parents were not taken in the study. Later, proportionate sampling was done to collect data from adolescents from each school.

### Data collection tools

A self-administered questionnaire, which was used to assess the status of depression, anxiety, stress, and self-esteem in adolescents and its relationship with parenting style, was divided into three sections. The first section includes information on socio-demographic characteristics, family characteristics, and academic characteristics. The second section includes questions to measure perceived parenting style using Parenting Style and Dimension Questionnaire – Short Version (PSDQ).[19] The Third section includes questions to measure the status of depression, anxiety, and stress among adolescents using the previously validated Nepali version of Depression, Anxiety and Stress Scale - 21 (DASS-21).[20] The Fourth section includes questions to measure self-esteem levels among adolescents using the Rosenberg Self-esteem Scale.[21] Parenting Style and Dimension Questionnaire – Short Version (PSDQ) was used to assess the adolescents’ perception of parenting style. The Short Version of the PSDQ consists of 32 items rated on a five-point Likert-type scale ranging from 1 (never) to 5 (always).[19] The Depression, Anxiety, and Stress Scale - 21 Items (DASS-21) is a set of three self-report scales designed to measure the emotional states of depression, anxiety, and stress. Each of the three DASS-21 scales contains 7 items, divided into subscales with similar content. Each item is rated on a 4-point Likert scale, ranging from 0 (indicating “did not apply to me at all”) to 3 (indicating “applied to me very much”). Scores for DAS were calculated by summing the scores for the relevant items and multiplying by two.[20] Rosenberg Self-esteem Scale, which was used to measure self-esteem in the study, is a 10-item scale that measures global self-worth by measuring both positive and negative feelings about the self. The scale is believed to be uni-dimensional. All items are answered using a 4-point Likert scale format ranging from strongly agree to strongly disagree.[21]

### Data collection procedure and technique

The data collection for the study was started on the mid September 2022 after the authorization of the ethical approval on the first week of September 2022. Data was collected after the permission from the municipal education division and individual secondary schools. The questionnaire was pretested among 58 school-going adolescents of Kathmandu Metropolitan. Self-administered questionnaires were distributed among the students, and orientation session was provided to students before data collection, and information on mental health issues was provided to students at the end of the session. Written informed consent was taken from all students prior to data collection whereas additional written parental consent was obtained from students below 18 years of age.

### Data analysis

The data entry form was created prior to data collection in IBM SPSS Statistics version 16 and was later entered, cleaned, and sorted for further analysis. Descriptive analysis was performed and frequency tables with percentage was generated for categorical variables and mean and standard deviation for continuous variables.

Binary logistic regression was performed to identify associated factors of symptoms of DAS. Firstly, we performed a univariate analysis in which each co-variate was modeled separately to determine the odds of DAS. Candidate variables for multivariable logistic regression were those whose univariate analysis yielded a p-value less than 0.05. A p-value of less than 0.05 was deemed statistically significant in multivariable logistic regression, and the strength of association was measured using the adjusted odds ratio (AOR) with a 95% confidence interval.

### Ethical approval and consent

All the activities associated with the study proceeded after acquiring the required authorization from the Manmohan Memorial Institute of Health Sciences’ Institutional Review Committee (MMIHS IRC 875; Ref 79/139; Date: 1 September 2022). Prior approval was sought from the municipality’s education sector and the school administration before data collection. Participants were also well-informed about the study. A written informed consent was obtained from the students before the data collection to ensure their willingness to participate and no identification was included in the questionnaire to ensure anonymity and confidentiality. Parental consent was obtained for students who were under the age of 18.

## Results

### Sociodemographic, family, and contextual characteristics of adolescents

The research questionaries were distributed among 583 school-going adolescents from 5 public and private schools each.

Table 1 demonstrates the demographic and family characteristics of the respondents. The mean age of the respondents was 15.41 ± 1.02 ranging from 10-18 years. Among 583 participants, more than half of the respondents (58.8%) were male in comparison to female respondents (40.8%). With regards to academic characteristics, about two-thirds of the respondents (64.5%) belonged to public schools and more than half (57.5%) were in grade 10. Five-sixth of the respondents (83%) were from Brahmin/Chettri ethnicity. Talking about the parent’s educational status, just over five-sixth of mothers (84.6%) were literate and the majority of the fathers (96.2%) were literate.

**Table 1.**
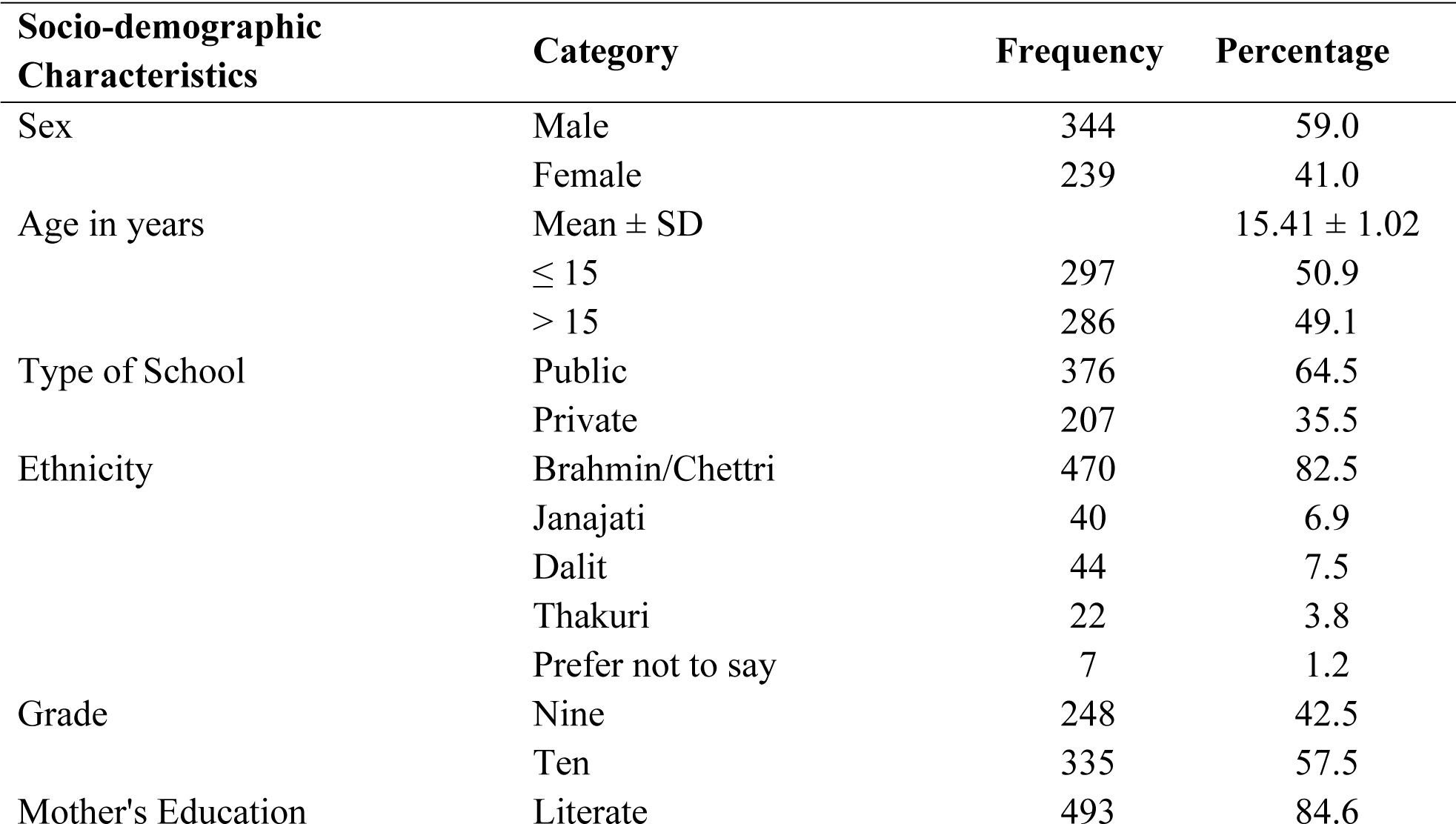
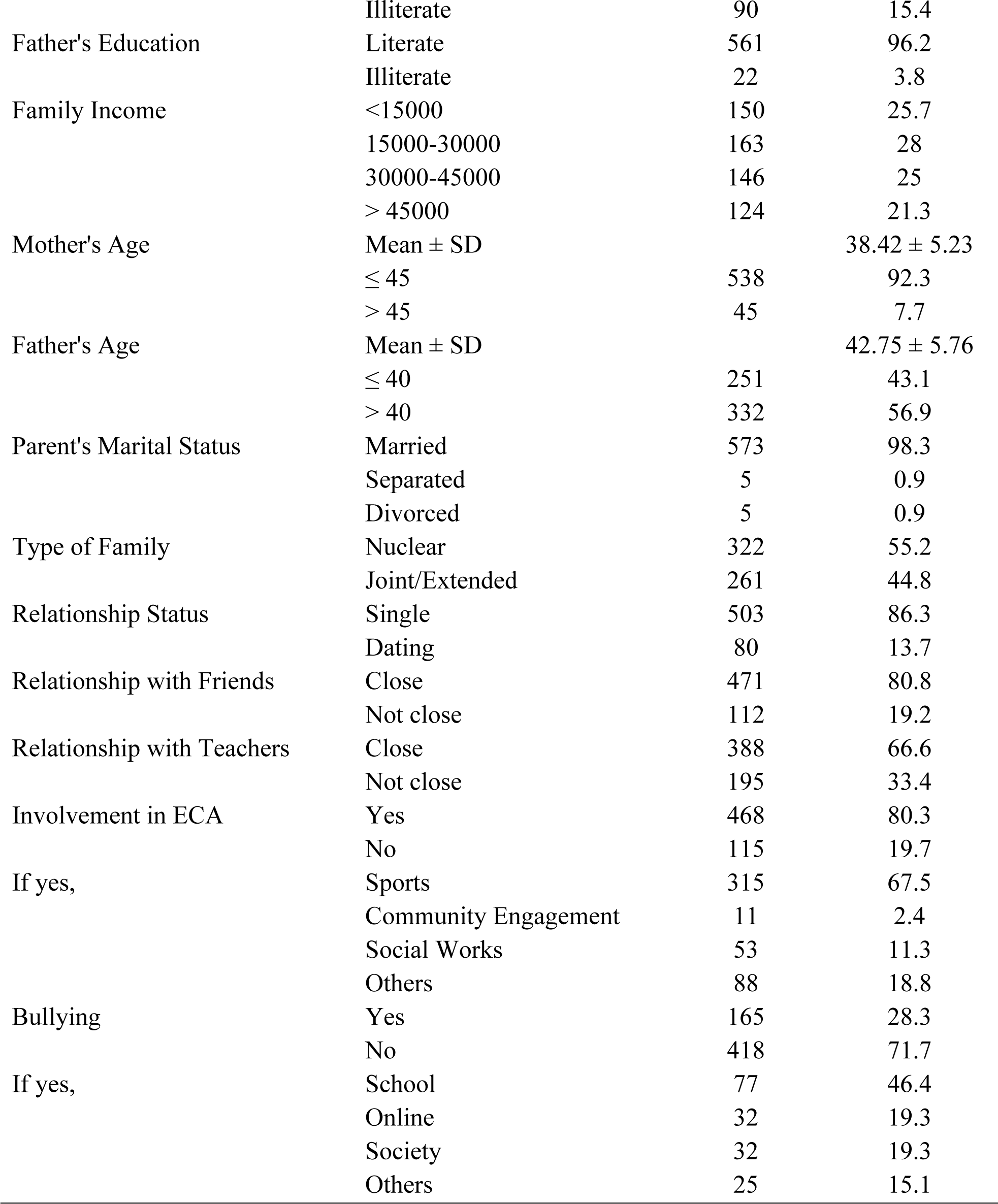
Socio-demographic characteristics of respondents.

Likewise, the majority of the students responded that their father as well as mother had attained a secondary level of education. Likewise, 15000-30000 was the majority of the income in most families, that is 28%. The majority of the parents (93%) were married while the remaining tiny fraction (0.9%) were separated and divorced respectively. More than half (55.2 %) of the respondents lived in the nuclear family.

It was noted that the majority of the adolescents (86.3%) were single and the majority of adolescents (80.8%) described their relationship with friends as close and two third (66.6%) of adolescents described their relationship with teachers as close. Talking about the involvement in extracurricular activities, it was found that the majority (80.3%) were involved in extracurricular activities (ECA) with just over two-thirds (67.5%) involvement in sports activity. Likewise, just over a quarter (28%) of the adolescents complained of being bullied, of which nearly half of the adolescents (46.4%) were bullied in the school environment.

### Perceived parenting style among adolescents

The results of perceived parenting style and its dimensions are shown in Table 2. It was found that the percentage of authoritative parenting style was 83.2%, authoritarian parenting style was 43.6% and permissive parenting style was 56.6%. The overall mean scores of authoritative, authoritarian, and permissive parenting styles were 4.16 ± 0.64, 2.18 ± 0.83, and 2.83 ± 0.84, respectively.

**Table 2.** Perceived parenting style among adolescents (n = 583)

| Parenting Style | Dimensions | Mean ± SD |
| --- | --- | --- |
| Authoritative | | $4.16 \pm 0.64$ |
| | Connection | $4.36 \pm 0.67$ |
| | Regulation | $4.35 \pm 0.70$ |
| | Autonomy | $3.77 \pm 0.95$ |
| Authoritarian | | $2.18 \pm 0.83$ |
| | Physical Coercion | $2.32 \pm 1.13$ |
| | Verbal Hostility | $2.45 \pm 1.05$ |
| | Non-Reasoning/Punitive | $1.77 \pm 0.89$ |
| Permissive | Indulgent | $2.83 \pm 0.84$ |

### Prevalence of depression, anxiety, and stress among adolescents

The prevalence of symptoms of depression, anxiety, and stress was found to be 37.39% (95% CI: 33.46%, 41.32%) 42.19% (95% CI: 38.18%, 46.19%), and 24.69% (95% CI: 21.18%, 28.19%) respectively. (Table 3)

**Table 3.** Prevalence of depression, anxiety, and stress among adolescents (n = 583) Depression Anxiety Stress.

| Level | Depression |  | Anxiety |  | Stress |  |
| --- | --- | --- | --- | --- | --- | --- |
|  | n | % | n | % | n | % |
| No | 365 | 62.6 | 337 | 57.8 | 439 | 75.3 |
| Mild | 83 | 14.2 | 49 | 8.4 | 49 | 8.4 |
| Moderate | 93 | 16.0 | 101 | 17.3 | 50 | 8.6 |
| severe | 19 | 3.3 | 41 | 7.0 | 34 | 5.8 |
| Extremely severe | 23 | 3.9 | 55 | 9.4 | 11 | 1.9 |

### Self-esteem among adolescents

The mean self-esteem score was 22.26 ± 3.56. About the majority of the adolescents (69.3%) showed high self-esteem while the remaining (30.7%) of the adolescents had low self-esteem.

### Association between parenting style and outcome variables

Table 4 represents the significant relationship between parenting style variables and depression, stress, anxiety, and self-esteem. It was found that authoritative parenting style has a low negative correlation with depression (r = -0.21) and anxiety variable (r = -0.20) and negligible negative correlation with stress (r = -0.19) and self-esteem variable (r = -0.16). Likewise, authoritarian parenting style showed a negligible positive correlation with DAS variables (r_d_ = 0.15, r_a_ = 0.13, r_s_= 0.08), while, low positive relation with self-esteem (r = 0.15). Similarly, the data revealed that permissive parenting style has a negligible positive correlation with DAS variables (r_d_ = 0.10, r_a_ = 0.06, r_s_= 0.11) and self-esteem (r = 0.07).

**Table 4.** Association between parenting style and outcome variables.

| Variables | Depression |  | Anxiety |  | Stress |  | Self-esteem |  |
| --- | --- | --- | --- | --- | --- | --- | --- | --- |
|  | r | p-value | r | p-value | r | p-value | r | p-value |
| Authoritative | -0.21 | <0.0001 | -0.20 | <0.0001 | -0.19 | 0.0001 | -0.16 | 0.0001 |
| Authoritarian | 0.15 | 0.0003 | 0.13 | 0.0019 | 0.08 | 0.0637 | 0.15 | 0.0004 |
| Permissive | 0.10 | 0.0132 | 0.06 | 0.1376 | 0.11 | 0.0060 | 0.07 | 0.1042 |

### Factors associated with depression

The results of multivariate logistic regression for the factors associated with depression are shown in Table 5. In the final model, the father’s age and bullying were found to be significantly associated with depression. Results showed that the odds of depression among adolescents having fathers aged >50 was 2.05 times (AOR=2.05, 95% CI 1.07 – 3.93) more likely than adolescents with fathers aged ≤ 50. Likewise, the odds of depression among adolescents who reported bullying was 2.42 times (AOR=2.42, 95% CI 1.64 – 3.55) more likely than in adolescents who didn’t report bullying.

**Table 5.** Factors affecting depression, anxiety, stress and self-esteem among adolescents.

| **Characteristics** | **Depression<sup>1</sup>**<br>**AOR, 95% CI** | **Anxiety<sup>2</sup>**<br>**AOR, 95% CI** | **Stress<sup>3</sup>**<br>**AOR, 95% CI** | **Self-esteem<sup>4</sup>**<br>**AOR, 95% CI** |
| --- | --- | --- | --- | --- |
| **Sex** |  |  |  |  |
| Male |  |  | Ref | 1.36 (0.94 – 1.98) |
| Female |  |  | 2.17 (1.44- 3.27) * | Ref |
| **Age** |  |  |  |  |
| ≤ 15 |  |  |  | Ref |
| > 15 |  |  |  | 1.88 (1.18 – 3.00) * |
| **Grade** |  |  |  |  |
| 9 |  | Ref | Ref |  |
| 10 |  | 1.81 (1.27 – 2.58) * | 1.73 (1.15 – 2.60) * |  |
| **Mother's education** |  |  |  |  |
| Illiterate | 1.50 (0.93 – 2.42) |  |  |  |
| Literate | Ref |  |  |  |
| **Father's education** |  |  |  |  |
| Illiterate |  |  |  | 3.79 (0.85 – 16.86) |
| Literate |  |  |  | Ref |
| **Father's age** |  |  |  |  |
| ≤ 45 | Ref |  |  |  |
| > 45 | 2.05 (1.07 – 3.93) * |  |  |  |
| **Relationship status** |  |  |  |  |
| Single |  |  |  | Ref |
| Dating |  |  |  | 1.63 (0.90 – 2.95) |
| **Relationship with friends** |  |  |  |  |
| Not close | 1.17 (0.76 – 1.84) | 1.99 (1.28 – 3.10) * | 1.71 (1.07 – 2.73) * | 2.68 (1.51 – 4.76) * |
| Close | Ref | Ref | Ref | Ref |
| **Relationship with teacher** |  |  |  |  |
| Not close | 1.33 (0.91 – 1.94) | 1.15 (0.79 – 1.68) | 1.95 (1.29 – 2.96) * | 1.08 (0.70 – 1.65) |
| Close | Ref | Ref | Ref | Ref |
| **Involvement in ECA** |  |  |  |  |
| No |  |  |  | 1.67 (1.01 – 2.76) * |
| Yes |  |  |  | Ref |
| **Bullying** |  |  |  |  |
| No | Ref | Ref | Ref | Ref |
| Yes | 2.42 (1.64 – 3.55) * | 2.26 (1.67 – 3.62) * | 1.95 (1.27 – 3.01) * | 2.37 (1.47 – 3.82) * |

<sup>1</sup>adjusted for education of mother, age of father, relationship with friends, relationship with teacher and bullying
<sup>2</sup>adjusted for grade of study, relationship with friends, relationship with teacher and bullying
<sup>3</sup>adjusted for sex of the respondent, grade of study, relationship status, relationship with teacher and bullying
<sup>4</sup>adjusted for age, grade of study, education of father, relationship with friends, relationship with teacher, involvement in ECA and bullying
OR: Odds Ratio
CI: Confidence Interval
Ref: Reference group
\*Statistically significant at $p < 0.05$

### Factors associated with anxiety

The results of multivariate logistic regression for the factors associated with anxiety are shown in Table 5. The variables that remained in the final model were grade, relationship with friends, and bullying. Grade (AOR=1.81, 95% CI 1.27 – 2.58), Relationship with friends (AOR=1.99, 95% CI 1.28 – 3.10), Relationship with teachers (COR=1.49, 95% CI 1.05 – 2.11) and bullying (AOR=2.26, 95% CI 1.67 – 3.62) was found to be significantly associated with symptoms of anxiety.

### Factors associated with stress

Factors associated with stress are presented in Table 5. Multivariate logistic regression between stress and selected independent variables showed that sex (AOR=0.46, 95% CI 0.31 – 0.69), grade (AOR=1.73, 95% CI 1.15 – 2.60), relationship with friends (AOR=1.71, 95% CI 1.07 – 2.73), relationship with teachers (AOR=1.95, 95% CI 1.29 – 2.96) and bullying (AOR=1.95, 95% CI 1.27 – 3.01) were significantly associated with symptoms of stress.

### Factors associated with self-esteem

The final model of multivariate logistic regression between self-esteem and selected independent variables showed that age (AOR=1.88, 95% CI 1.18 – 3.00), relationship with friends (AOR=2.68, 95% CI 1.51 – 4.76), involvement in ECA (AOR=1.67, 95% CI 1.01 – 2.76) and bullying (AOR=2.37, 95% CI 1.47 – 3.82) were significantly associated, as displayed in Table 5.

## Discussion

The present study elucidates the intricate associations between parenting styles and various psychological outcomes, including depression, anxiety, stress, and self-esteem among adolescents. Our findings underscore the nuanced influence of parenting approaches, with the authoritative parenting style exhibiting a low negative correlation with DAS and self-esteem while the authoritarian and permissive parenting style showed a negligible positive correlation across these psychological dimensions.

These results are consistent with the data obtained in research by Sanjeevan, et. al which indicated that authoritative parenting style correlated with a lower level of DAS while neglectful parenting style with a higher level of DAS.[14] A cross-sectional study on US adolescents revealed parental care and control were associated with mental health disorders among adolescents with involved parenting having a positive role and neglectful parenting having the negative role.[22] Likewise, Mishra et. al demonstrated a significant relationship between anxiety and parents adopting an authoritative parenting style and has shown an increase in social anxiety in adolescents with authoritative parents.[23] The current study found that authoritative parenting style was significantly associated with depression. This outcome is corroborated by that of Prativa et. al which indicated a negative correlation between authoritative parenting style and depression in adolescents that is more authoritative parenting style is associated with less depression in adolescents.[15] Similar results were found in the study conducted in Japan in later adults [24] and in Indonesia in high school students.[25]

Research findings showed a negative association of self-esteem with authoritative parenting style and a positive association with authoritarian and permissive parenting styles. This differs from the existing study conducted on the urban high school students of Nepal by Banstola et al. which reported a positive association between parental authoritativeness and self-esteem.[12] This discrepancy might be due to differences in the study setting and population. Likewise, a study in Indonesia showed a beneficial effect of authoritative parents on self-esteem and a negative effect of authoritarian parents on self-esteem, which differs from the research findings.[25] Research study by Milevsky et. al revealed the supportive role of an authoritative parenting style in increasing self-esteem.[11] The possible explanation for these results may be due to the nature of parenting style. It has been evident that an authoritative parenting style is characterized by parents who tend to display love and warmth, and high responsiveness towards the children. This reason might have resulted in a decrease in the symptoms of DAS among school-going adolescents.[22] However, the authoritative parenting style showed a negative correlation with self-esteem which resulted in a decrease in self-esteem. The result might be explained by the fact that the control nature of authoritative parents might increase the high expectations towards children as a result this will decrease self-esteem in adolescents. Likewise, authoritarian and permissive parenting styles showed negative association with DAS and self-esteem. This relationship may partly be explained by the fact that authoritarian parents try to keep their children in the controlled environment pressurizing their children which will increase DAS.[22,26] Likewise, permissive parents are less involved with their children which might have increased DAS.[26,27]

Study results revealed the prevalence of depression, anxiety, and stress to be 37.39%, 42.19%, and 24.69% respectively. Existing research conducted by Gautam et. al in the rural setting of Nepal showed about one-quarter of the high school students that is 27% showed depressive symptoms[28] which is lower than the study. Likewise, Sharma et. al research conducted among public schools of Kathmandu Valley reported the higher prevalence of depressive symptoms which is 41.6%.[29] Similarly, more than half that is 56.9% of students showed symptoms of anxiety and more than 1/4^th^ of students that is 27.5% showed symptoms of stress corroborating research findings.[29] Another study conducted by Bhandari et. al revealed that roughly one out of every two students (46.5%) experienced symptoms of anxiety.[30] Likewise, the study by Karki et. al revealed more than half (56.5%) of the high school students of Kathmandu showed symptoms of depression and a similar proportion (55.6%) showed anxiety symptoms, and about 1/4^th^ of the students showed stress symptoms that is 32.9%.[18] These findings are inconsistent with current research. The prevalence of depression was found high as compared to study conducted in the rural setting of Nepal. One of the possible explanations for this could be that data was collected at the very exam season. During this period students may have struggled academically and displayed symptoms of depression, anxiety and stress. Likewise, majority of the students in the study were grade 10 students, who were under constant pressure for approaching SEE exams. Taking into account academic characteristics of the adolescents, grades showed significant association with stress. This was in line with a study conducted in India[31] and Bangladesh.[32] Among South Asian countries, the prevalence of depression reported by our study is lower than the studies conducted in India, Bangladesh, and China.[33–36] On the contrary, the prevalence of anxiety as reported by the current study is in line with the studies conducted in China and Vietnam, but higher than that of Sri Lanka and lower than that of India.[33,35,37,38] Likewise, the prevalence of stress is comparable with the study conducted in Manipur, India but lower than that of a similar study conducted in Chandigarh, India.[34,39] These findings suggested the increasing burden of DAS among school-going adolescents in South Asia. The mean self-esteem score in this study was found to be 22.26 ± 3.56. The study has shown that the majority of the adolescents (69.3%) showed high self-esteem scores. This was in line with the study conducted in Nigeria (24.0 ± 3.3).[40]

However, this differs from the research findings of Banstola et. al (16.59 ± 0.16)[12] and study in Iran (38.49 ± 6.55).[41] The possible reason for this might be the difference in study setting and population.

In the current study, it was found that female adolescents were more likely to suffer from symptoms of stress than male adolescents. The findings of the study are consistent with the research of Sharma et. al which showed higher symptoms of stress in females.[18] This is inconsistent with the findings of the previous research studies.[37,42–45] The possible explanation must be the difference in parenting between male and female adolescents which can result in differences in the mental status of the adolescents. Likewise, females are more likely to be exposed to the family burden and social stigma. Similarly, the current study showed higher self-esteem scores in male adolescents than in female adolescents. These results agree with the findings of a study by Banstola et. al.[12] Likewise, study in Iran showed higher self-esteem in male adolescents than female adolescents.[41] On the contrary, another study in Iran reported higher mean scores of girls than boys.[16]

This study found the risk of depression was higher in adolescents with mothers without any formal education. The possible explanation might be illiterate mothers don’t have a stronger knowledge of the child’s mental health status and how to react to the child’s aspirations and demands which in turn can result in early signs of depression in the child. This differs from the findings of the study conducted by Gautam et. al.[28] Likewise, research by Bhattarai et. al revealed depression was higher in adolescents with illiterate parents.[46] Similarly, adolescents with illiterate fathers showed low self-esteem among adolescents. This was in accordance with the research findings of Alami et. al.[41] The possible explanation might be that illiterate fathers mightn’t have the knowledge to boost of confidence of their children. However, educated fathers play an important role in robust their child decision-making skills and boosting their morale.

Relationships with friends and teachers showed an association with DAS and self-esteem. It was found that the odds of depression, anxiety, stress, and low self-esteem among adolescents having no close relationship with friends was 1.52 times, 1.99 times, 1.71 times, and 2.68 times more likely than adolescents with close relationships with friends respectively. This was in accordance with the findings of the study conducted by Gautam et. al which showed those students who have poor relationships with friends were 2.4 times more likely to be at risk of depression than those students who have a good relationship with their friends.[28] Likewise, it was found that poor relationships with teachers resulted in adolescents experiencing 1.4 times more depressive symptoms.[28] Similar studies conducted among Chinese students showed having average relationships with classmates (AOR=2.82) or poor relationships with classmates (AOR=1.60) was a risk factor for having depressive symptoms.[47] Having not close relationship might have resulted in a lack of social support which eventually resulted in early signs of DAS and low self-esteem. A similar study on high school students showed a significant association between perceived social support and level of depression.[46] Likewise, anxiety symptoms were 1.73 times more likely in adolescents with poor relationships with classmates or friends[48], which was consistent with the findings of the current study. A possible explanation might be the fact that a lack of social support results in a lack of confidence to tackle issues and worry about small problems.

Bullying was also found to be significant among adolescents and was associated with increased levels of DAS and a decrease in self-esteem among adolescents. Likewise, the odds of depression, anxiety, stress, and low self-esteem among adolescents who reported bullying were 2.42 times, 2.17 times, 1.95 times, and 2.37 times more likely than in adolescents who didn’t report bullying respectively. It is consistent with the research findings of Karki et. al which showed the symptoms of depression were 2.84 times more likely among the students who were bullied electronically, symptoms of anxiety and stress were 1.36 and 1.48 times more likely in adolescents who were bullied in school property respectively.[18] Likewise, a similar study in Chinese high school students revealed that bullying victimization results in an increased level of social anxiety with the mediating role of self-esteem.[49] Similarly, another study showed consistent findings which confirmed that bullying victims have higher depressive symptoms, lower self-esteem, and higher suicide tendencies than non-victims.[50] Existing research on students also confirmed the significant association between stress and bullying.[51] The students who were bullied repeatedly experience abuse and threats which exacerbates the feeling of fear and loneliness. This causes emotional distress in adolescents, which leads to increased DAS symptoms and low self-esteem.

The findings of the study should be interpreted in light of its limitations. This study took the perspective of adolescents on the perceived parenting style. This perception might differ among the parents. Future studies should try to include the perspectives of both parents and children. Likewise, maternal and parental parenting style was found to be different and can interfere with the findings so future studies should consider taking the perception of parenting style from each parent.

## Conclusion

This study planned to assess depression, anxiety, stress, and self-esteem in adolescents and its relationship with parenting style in Bheemdatt Municipality, Kanchanpur. About one-third of adolescents showed symptoms of depression, almost half of the adolescents showed symptoms of anxiety, and about one-fourth of adolescents showed symptoms of stress in Bheemdatt Municipality. Likewise, the majority of the adolescents showed high self-esteem and about one-third of adolescents showed low self-esteem. Importantly, authoritative parenting style positively predicted the DAS and negatively predicted self-esteem while authoritarian and permissive parenting style negatively predicted the DAS and positively predicted the self-esteem. This shows the important role of parenting behavior in the prediction of the mental health status of adolescents with positive parenting behavior showing a positive impact on the behavior of the child. Study covariates such as sex, grade, parent’s age, and parent’s education showed strong effects on the outcome of the research. Likewise, the social support system of the adolescents was found to be a strong predictor of DAS and self-esteem. Thus, improving adolescents’ perceived level of social support may alleviate the mental health problems of adolescents. Similarly, students who were bullied showed a higher prevalence of DAS and low self-esteem. Therefore, prevention and control activities such as school-based counseling focusing on bullying will help to reduce mental health distress among adolescents. Hence, it is concluded that parents play an influential role in promoting the mental health of their adolescent children with the supporting role of study covariates and social support systems.

## Supporting information

**S1 Table. Study variables and tables.** This is the study variables used in the study and tables from further analysis done in the study.

**S1 Tool. Study tool.** English Questionnaire

**S1 Data. Study dataset.** Final version of study dataset

**S1 Checklist. STROBE Statement.** This is the Strengthening the Reporting of Observational Studies in Epidemiology (STROBE) Statement guidelines for reporting observational studies

## Data Availability

All data are available in the Supporting information files.

10.6084/m9.figshare.26535427

## Acknowledgment

Foremost, we express our sincere gratitude towards the students for their cooperation and support during the study. Special thanks to Bheemdatt municipality for granting permission to conduct the study. Grateful to the school administration and teachers for their assistance during data collection. Above all sincere gratitude towards and Department of Public Health, Manmohan Memorial Institute of Health Sciences for their unwavering leadership and assistance.

## Author contributions

**Conceptualization:** Anjali Bhatt, Rabina Khadka

**Data curation:** Anjali Bhatt

**Formal analysis:** Anjali Bhatt, Milan Thapa

**Investigation:** Anjali Bhatt

**Methodology:** Anjali Bhatt, Rabina Khadka

**Project administration:** Anjali Bhatt

**Supervision:** Rabina Khadka, Durga Khadka Mishra

**Visualization:** Anjali Bhatt

**Writing– original draft:** Anjali Bhatt, Rabina Khadka

**Writing– review & editing:** Anjali Bhatt, Rabina Khadka, Milan Thapa, Anusha Sharma, Manoj Joshi, Durga Khadka Mishra

